# Effects of voluntary event cancellation and school closure as countermeasures against COVID-19 outbreak in Japan

**DOI:** 10.1101/2020.03.19.20037945

**Authors:** Yoshiyuki Sugishita, Junko Kurita, Tamie Sugawara, Yasushi Ohkusa

**Affiliations:** National Institute of Infectious Diseases, Tokyo, Japan; Department of Nursing Collage of Nursing, Tokiwa University, Ibaraki, Japan

**Keywords:** COVID-19, voluntary event cancellation, outbreak in Japan, SIR model, asymptomatic case

## Abstract

**Background:** To control the COVID-19 outbreak in Japan, sports and entertainment events were canceled and schools were closed throughout Japan from February 26 through March 19. That policy has been designated as voluntary event cancellation and school closure (VECSC).

**Object:** This study assesses VECSC effectiveness based on predicted outcomes. Method: A simple susceptible–infected–recovery model was applied to data of patients with symptoms in Japan during January 14 through March 25. The respective reproduction numbers were estimated before VECSC (R), during VECSC (R_e_), and after VECSC (R_a_).

**Results:** Results suggest R before VECSC as 1.987 [1.908, 2.055], R_e_ during VECSC as 1.122 [0.980, 1.260], and R_a_ after VECSC as 3.086 [2.529, 3.739].

**Discussion and Conclusion:** Results demonstrated that VECSC can reduce COVID-19 infectiousness considerably, but the value of R rose to exceed 2.5 after VECSC.

## Introduction

The initial case of COVID-19 in Japan was a patient returning from Wuhan, China who showed symptoms on January 3, 2020. Subsequently, as of April 6, 2020, the Ministry of Labour, Health and Welfare (MLHW) in Japan reported 3,906 cases in Japan, including asymptomatic cases but excluding those from a large cruise ship: the Diamond Princess [1].

Sports and entertainment events were canceled in Japan for two weeks from February 26 through March 11 in compliance with a government advisory. At that time, it was advised that small business and private meetings be cancelled voluntarily: the measure was not enacted as a law or enforced by authorities. Therefore, people were not arrested or cited even if they did not comply with this government advisory [2]; the effort depended entirely on voluntary compliance of individuals. Moreover, this was the first measure necessitating voluntary event cancellation. Those reasons complicate the *ex ante* prediction of the proportion of events that were cancelled and the extent to which contact among people was reduced. Moreover, as announced March 3, almost all schools were closed from the middle of March through spring vacation (early April) as a measure to control the spread of COVID-19. Even though young people can be infected and can transmit the virus to adults, school closure effects were questionable *ex ante* because schoolchildren were not regarded as the most susceptible age class for COVID-19 [3–5]. Therefore, these policies must be evaluated *ex post*. The policies are known collectively as voluntary event cancellation and school closure (VECSC). After the VECSC period, VECSC ceased, but cancellation of large events was continued.

If the reproduction number under VECSC (R_e_) was less than one, then the outbreak can be expected to have been contained. Alternatively, even if R_e_ was markedly less than R_0_ but greater than one, one could have expected instead that the outbreak would be exacerbated.

This study was conducted to evaluate VECSC based on the epidemic curve because no evidence exists to reflect the number of cancelled events. The obtained result might be expected to contribute to a government decision to implement VECSC as a countermeasure in Japan.

## Method

We applied a simple deterministic susceptible–infected–recovery(SIR) model [6] to the epidemic curve in Japan for its population of 120 million. We assume an incubation period following the empirical distribution from the early stage of the outbreak. A person on the *i*-th day of incubation will move to a symptomatic or an asymptomatic state with probability of *p(i)/Σ_j_=_i ^L^_p(j)*, where *p*(*i*) represents the proportion of the *i*-th day on the empirical distribution of the incubation period and *L* denotes the maximum length of the incubation period. Alternatively, a person moves to the *i+*1 day in the incubation state with one minus *p(i)/Σ_j=i_^L^p(j)*.

Symptomatic and asymptomatic states were assumed to continue for seven days, with subsequent transition to a recovery state with probability of one [7]. This model ignores outcomes and necessary medical resources: states of death and hospitalization were not incorporated into the model.

Asymptomatic cases are not observable unless complete laboratory-based surveillance is performed. One exceptional study indicated them as 3/23 from a sample of elderly people [7]. We checked its robustness for the proportion of asymptomatic cases assuming a ratio of 4/19 from experiences of Japanese residents of Wuhan up to the outbreak [8].

Infectivity by severely infected patients and by mild patients was assumed to be equal. Moreover, we assumed that asymptomatic cases have the same power of infectivity as symptomatic cases [7]. That is merely an assumption. Therefore, we verified its robustness through sensitivity analysis of the infectiousness of asymptomatic cases, such as 50% assumed for simulation studies for influenza [9–13]. The distribution of infectiousness in symptomatic and asymptomatic cases was assumed to be 30% on the onset day, 20% on the following day, and 10% for the subsequent five days [7]. Regarding its robustness, we also applied a uniform distribution for a week.

Because VECSC was conducted during February 27 – March 19, we divided the data period into three periods of those before VECSC, during VECSC, and after VECSC, with corresponding reproduction numbers represented as R_0_, R_e_, and R_a_. We modeled the change in the reproduction number given the prior day’s reported number of persons susceptible, incubation, symptomatic, asymptomatic, and in a recovered state.

For this study, R_0_ was defined as the number of newly infected persons from one person infected over the infectious period when all persons were susceptible with no countermeasures. In addition, R_e_ and R_a_ were defined, respectively, as the numbers of newly infected persons from one person infected over the infectious period when all persons were susceptible under VECSC or during a time of countermeasures after the VECSC period. For estimation, the number of the newly infected persons was calculated using R_0_ (R_e_ or R_a_) × the proportion of susceptible persons among the population × the number of persons with infectiousness weighted by their degree of infectiousness.

The values of R_0_, R_e_, and R_a_ were sought to fit the data to minimize the sum of absolute values of discrepancies among the bootstrapped epidemic curve and the fitted values. The estimated distribution of three reproduction numbers was calculated using 10,000 iterations of bootstrapping for empirical distribution of the data for symptomatic patients. In this sense, although this model is deterministic at each bootstrapping iteration, its range can be estimated through 10,000 iterations.

The bootstrapping procedure we used with fully replicated bootstrapping for a fixed number of initial cases. There were *N* patients in the data, with numbering of the patients from the initial case to the last case. Initially, no patient was on the bootstrapped epidemic curve. If a random variable drawn from a uniform distribution of (0,1) was *t* included in the internal [*i*/(*N*-1), (*i+*1)/(*N*-1)], then we added one to the onset date of *i*+1th patient to the bootstrapped epidemic curve. We replicated this procedure *N*-1 times. Thereby, we obtain the bootstrapped epidemic curve with *N*-1 patients. Finally, we added the initial patient, for whom the onset date was January 14, to the bootstrapped epidemic curve. Consequently, we obtained a bootstrapped epidemic curve with *N* patients starting from January 14.

We estimated the curve sequentially as follows. First, we estimated R_0_ as the best fit to bootstrapped data for the pre-VECSC period. Then, based on the obtained R_0_ and the course of the outbreak before the VECSC period, we estimated R_e_ as the best fit to bootstrapped data in the VECSC period. Finally, based on the obtained values of R_0_ and R_e_, we estimated R_a_ as the best fit to bootstrapped data for the post-VECSC period. In each step, reproduction numbers were grid searched in the interval of (0,10) by 0.001.

We conducted some sensitivity analyses for one way in addition to the base case as explained above: the infectiousness power of asymptomatic cases represented 50% of symptomatic cases; the proportion of asymptomatic cases was 4/19 as symptomatic cases; infectiousness and infectious patterns showed a uniform distribution for one week. We also estimated the parameters by minimizing the sum of the squared residuals or maximizing Poisson likelihood instead of the difference in absolute values and using the fitted incubation period as a log normal distribution instead of the empirical distribution.

Moreover, we examined the possibility of infection during two days before onset. Its probability can be represented as *Σ*_*i* 2_^L-1^*p(i)*/*Σ*_*j=i*_^L^*p(j) x*(*i*-1,*t*-1) + (1-*p(i)*/*Σ *p(j)*) p(i+1)/*Σ*_j=i_+1^L^p(j) x*(*i*-1,*t*-2)*+ x*(*L-1,t*-1), where *x*(*i,t*) represents the number of persons at the *i-*th incubation period on day *t*. They were assumed to have the same infectiousness in the first two days of symptomatic period. To meet the definition of the reproduction number, infectiousness in the incubation and (a) symptomatic period were adjusted such that the sum of infectiousness from two days before onset to one week following onset would be one.

We also examined incorporation of the imported cases into the model. They were entered into the model as newly symptomatic cases with no history of incubation period when the imported cases were reported. The parameters in this model were estimated as minimizing the sum of the absolute difference among the number of patients including the imported cases and prediction.

We adopted 5% as the level for which we inferred significance. We used Matlab 2014a to code the model as explained above.

### Data source

The numbers of symptomatic patients during January 14 – March 25 were published by the MHLW [1] and were reported publicly from 46 prefectures and some cities

(https://www.pref.okinawa.lg.jp/site/hoken/chiikihoken/kekkaku/covid19_hasseijoukyou.html;

https://www.pref.kagoshima.jp/ae06/kenko-fukushi/kenko-iryo/kansen/kansensho/coronavirus.html;

https://www.pref.miyazaki.lg.jp/kansensho-taisaku/covid-19/hassei_list.html;

https://www.pref.oita.jp/site/covid19-oita/covid19-pcr.html;

https://www.pref.kumamoto.jp/kiji_32300.html;

https://www.pref.nagasaki.jp/bunrui/hukushi-hoken/kansensho/corona_nagasaki/corona_nagasaki_shousai/;

https://www.pref.saga.lg.jp/kiji00373220/index.html;

https://www.pref.fukuoka.lg.jp/contents/covid19-hassei.html;

https://www.city.fukuoka.lg.jp/hofuku/hokenyobo/health/kansen/cohs.html;

https://www.pref.kochi.lg.jp/soshiki/130401/2020022900049.html;

https://www.pref.ehime.jp/h25500/kansen/covid19.html#kansensha;

https://www.pref.kagawa.lg.jp/content/dir1/dir1_6/dir1_6_2/wt5q49200131182439.shtml#outbreak;

https://www.pref.yamaguchi.lg.jp/cms/a10000/korona2020/202004240002.html;

https://www.pref.hiroshima.lg.jp/soshiki/57/bukan-coronavirus.html;

https://www.pref.okayama.jp/page/645925.html#kennaijoukyou;

https://www.pref.shimane.lg.jp/bousai_info/bousai/kikikanri/shingata_taisaku/new_coronavirus_portal.html;

https://www.pref.tottori.lg.jp/291425.htm;

https://www.pref.wakayama.lg.jp/prefg/041200/d00203387.html;

http://www.pref.nara.jp/module/1356.htm#moduleid1356;

https://web.pref.hyogo.lg.jp/kk03/corona_hasseijyokyo.html;

http://www.pref.osaka.lg.jp/hodo/index.php?HST_TITLE1=%83R%83%8D%83i&SEARCH_NUM=10&searchFlg=%8C%9F%81@%8D%F5&site=fumin;

https://www.pref.kyoto.jp/kentai/news/novelcoronavirus.html#F;

https://www.pref.shiga.lg.jp/ippan/kenkouiryouhukushi/yakuzi/310735.html;

https://www.pref.mie.lg.jp/YAKUMUS/HP/m0068000066.htm;

https://www.pref.aichi.jp/site/covid19-aichi/corona-kisya.html;

http://www.city.nagoya.jp/kenkofukushi/page/0000126920.html;

https://www.pref.shizuoka.jp/kinkyu/covid-19-tyuumokujouhou.html;

https://www.pref.gifu.lg.jp/kinkyu-juyo-joho/shingata_corona_kansendoko.html;

https://www.pref.nagano.lg.jp/hoken-shippei/kenko/kenko/kansensho/joho/corona-doko.html;

https://www.pref.yamanashi.jp/koucho/coronavirus/info_coronavirus_prevention.html;

https://www.pref.fukui.lg.jp/doc/kenkou/corona/jyoukyou.html;

https://www.pref.ishikawa.lg.jp/kansen/coronakennai.html;

http://www.pref.toyama.jp/cms_sec/1205/kj00021798.html;

https://www.pref.niigata.lg.jp/sec/kenko/covid19.html;

https://www.pref.kanagawa.jp/docs/ga4/bukanshi/occurrence_06.html;

https://www.bousai.metro.tokyo.lg.jp/taisaku/saigai/1007261/index.html;

https://www.pref.chiba.lg.jp/shippei/press/2019/ncov-index.html;

https://www.pref.saitama.lg.jp/a0701/shingatacoronavirus.html;

https://www.pref.gunma.jp/07/z87g_00016.html;

http://www.pref.tochigi.lg.jp/e04/welfare/hoken-eisei/kansen/hp/coronakensahasseijyoukyou.html; https://www.pref.ibaraki.jp/1saigai/2019-ncov/hassei.html;

https://www.pref.fukushima.lg.jp/sec/21045c/fukushima-hasseijyoukyou.html;

https://www.pref.yamagata.jp/ou/bosai/020072/kochibou/coronavirus/coronavirus.html

#kensa;

https://www.city.yamagata-yamagata.lg.jp/kakuka/kenkoiryo/kenkozoshin/sogo/kansensyou/pd0409180023.html;

https://www.pref.akita.lg.jp/pages/archive/47957;

https://www.pref.miyagi.jp/site/covid-19/02.html;

https://www.pref.iwate.jp/kurashikankyou/iryou/covid19/index.html;

https://www.pref.aomori.lg.jp/welfare/health/wuhan-novel-coronavirus2020.html;

http://www.pref.hokkaido.lg.jp/hf/kth/kak/hasseijoukyou.htm#4/12) as of April 6.

During this period, 1516 cases were recorded with onset dates. We excluded imported cases and cases representing infected persons from the Diamond Princess because they were presumed not to have been community-acquired in Japan.

### Ethical considerations

All information used for this study has been published elsewhere [1]. There is therefore no ethical issue related to this study.

## Results

Figure 1 depicts the empirical distribution of the incubation period among 91 cases for which the exposed date and onset date were published by MHLW. Its mode was six days. The average was 6.6 days. The maximum incubation period length was 17 days.

**Figure 1:**
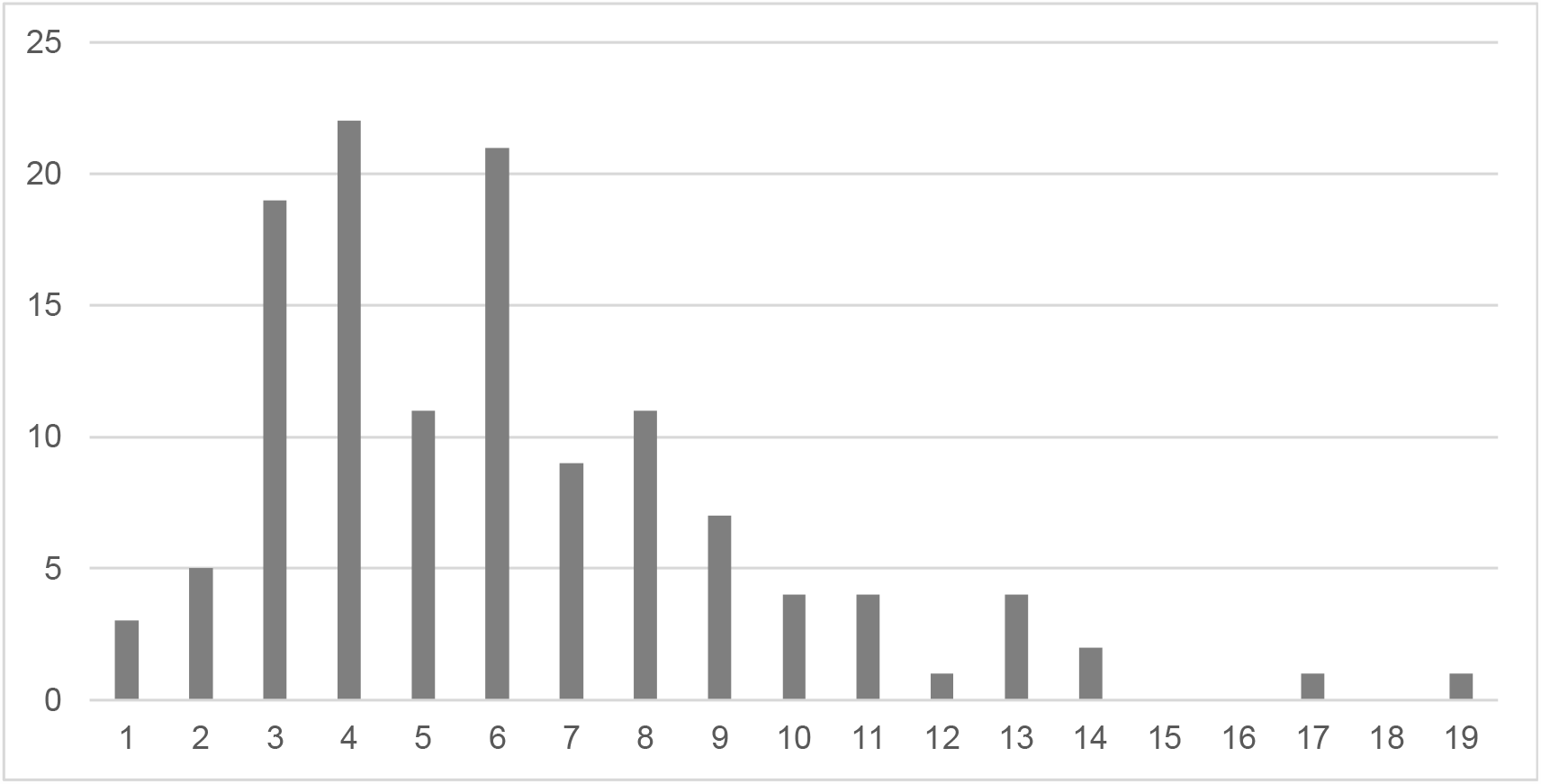
Empirical distribution of incubation period published by Ministry of Labour, Health and Welfare, Japan Notes: Bars shows distribution of incubation period among 91 cases whose exposed date and onset date were published by Ministry of Labour, Health and Welfare, Japan. Because the patients whose incubation was longer than 14 days were just 4%, they were included as bars on 14.

The value of R_0_ before introduction of VECSC was estimated as 2.534. Its range was [2.449, 2.598]. However, R_e_ during the VECSC period was estimated as 1.077 [0.948, 1.228]. After VECSC, R_a_ was estimated as 4.455 [3.615, 5.255]. The null hypothesis that R_0,_ R_e_, and R_a_ were equal was therefore rejected.

Figure 2 depicts the observed epidemic curve and the predicted epidemic curve based on the estimated R_0_, R_e_, and R_a_. It showed goodness of fit that was pretty good. The small bump around February 26 reflects overshooting caused by the sharp decline of the reproduction number from R_0_ to R_e_.

**Figure 2:**
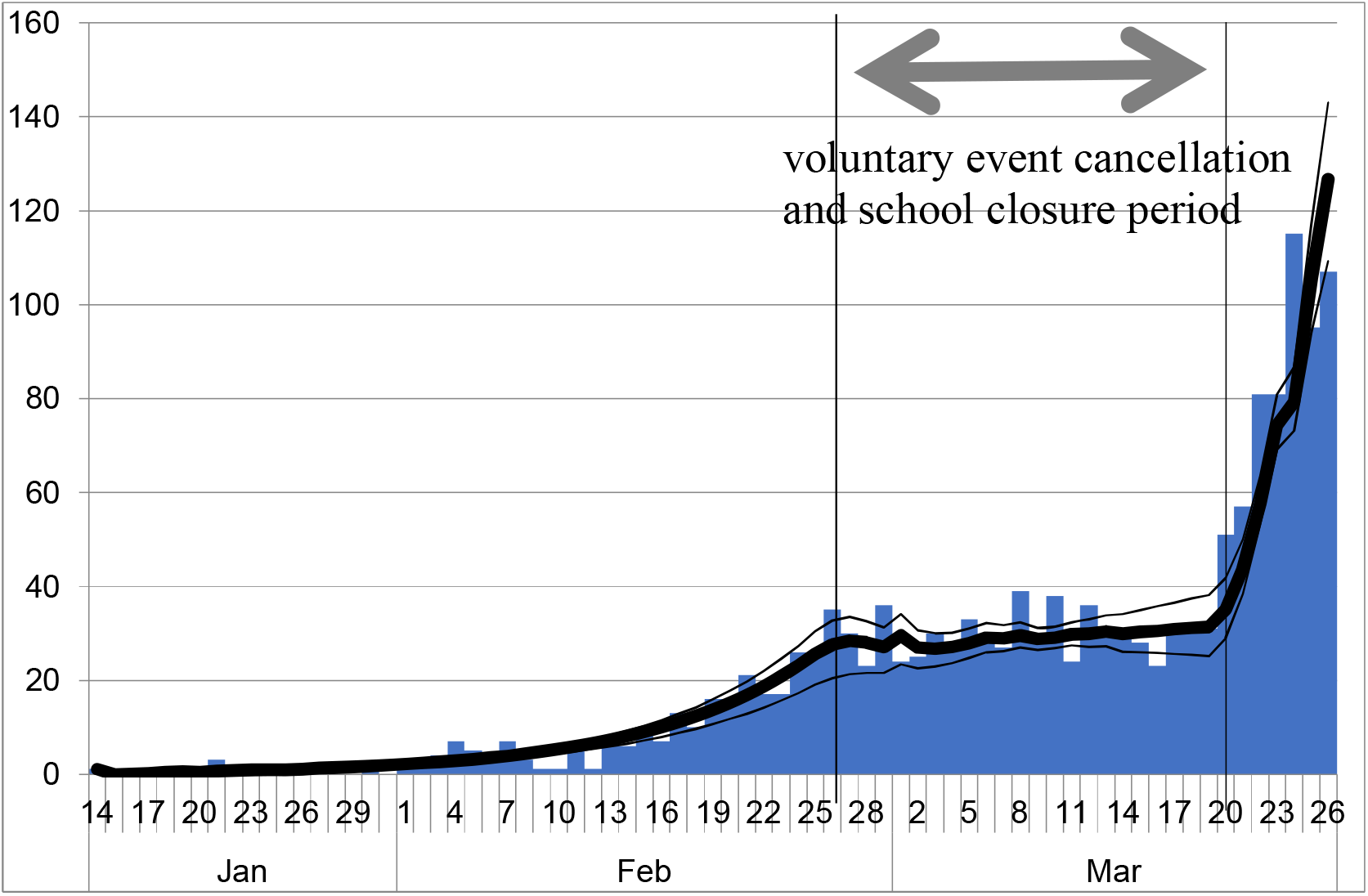
Observed epidemic curve of COVID-19 patients and predicted epidemic curve from the model based on the estimated reproduction number Note: Bar indicates the observed epidemic curve and bold line indicates the predicted line based on the estimated reproduction numbers. Thin lines indicate range of the best fit line at each bootstrapping iteration. Between two vertical lines indicates voluntary event cancellation and school closure period.

One-way sensitivity analysis revealed that if the infectiousness of asymptomatic cases was 50% of symptomatic cases instead of 100%, then R_0_ would be estimated as 2.886 [2.801, 2.970], R_e_ would be 1.203 [1.059, 1.343], and R_a_ would be 4.985 [4.134, 5.893].

If the asymptomatic cases were 4/19 as symptomatic cases instead of 3/32, then R_0_ would be estimated as 2.0492 [2.403, 2.557], R_e_ would be 1.091 [0.972, 1.237], and R_a_ would be 4.428 [3.627, 5.049]. If the infectious pattern formed a uniform distribution for a week instead of the base case, then R_0_ would be estimated as 2534 [2.449, 2.598], R_e_ would be 1.077 [0.948, 1.228], and R_a_ would be 4.455 [3.615, 5.255]. If we introduce infectiousness in the incubation period, then R_0_ would be estimated as 2.877 [2.765, 2.980], R_e_ would be 1.278 [1.085, 1.478], and R_a_ would be 4.687 [3.621, 5.000].

When we estimate the parameters by minimizing the sum of the squared residuals instead of the absolute difference, R_0_ was estimated as 2.391 [2.505, 2.609], R_e_ was 1.124 [0.935, 1.295], and R_a_ was 4.409 [3.168, 5.341]. In the case of using Poisson likelihood as evaluation, the parameters were estimated as 2.007 [1.957, 2.058], 1.141 [1.030, 1.237], and 3.255 [2.697, 3.922]. However, when we estimate the parameters using a log normal distribution with mean of 1.66 and standard deviation of 0.558 truncated as 17 days as incubation period instead of the empirical distribution, R_0_ was estimated as 2.2660 [2.179, 2.838], R_e_ was 1.116 [0.970, 1.277], and R_a_ was 3.574 [2.897, 4.338].

However, we found 217 imported cases in this period, representing 14.3% of the community acquired patients. When we incorporated the imported cases into the model, R_0_ was estimated as 2.502 [2.412, 2.577], R_e_ was 1.113 [0.978, 1.298], and R_a_ was 4.311 [3.603, 5.103]. Because the ranges of R_e_ and R_a_ were overlapped in all cases, these factors might not greatly affect both parameters, at least in the region with parameters and distribution that are considered reasonable. The estimated R_0_ was the most sensitive. When using Poisson likelihood for evaluation, R_0_ was significantly lower than in the base case. Moreover, infectiousness before onset raised R_0_ significantly. In other scenarios, the ranges of R_0_ were overlapped.

## Discussion

We applied a simple deterministic SIR model including asymptomatic cases. A deterministic model might be simpler but more appropriate than a stochastic model for the present study including changing parameters and policy evaluation [14].

An earlier study [15–17] estimated R_0_ for COVID-19 as 2.24–3.58 in Wuhan. Our R_0_ obtained for the period before VECSC was similar. However, an earlier study [18] found the average number of persons who were secondarily infected with COVID-19 in Japan as 0.6 before the VECSC period, with 80% of patients found to have infected no one. If the average number of secondarily infected persons was less than one, then the outbreak might stall on its own. Given such an eventuality, no countermeasures would have been required. However, based on results of a study uploaded to the site of Japanese Public Health Association, health authorities in the public health sector or local government were urged to conduct contact tracing to detect clusters [19]. Because only a few patients infected other people, as found from this study, then the outbreak might be controlled if clusters could be suppressed.

Infectiousness two days before symptom onset has been described in the literature [7, 20]. We applied it as sensitivity analysis. It showed slightly higher reproduction numbers than the base model, although the estimated R_e_ and R_a_ were found to have a non-significant difference from the base case because a longer period of infectiousness implies fewer infected people on a day when the reproduction number does not change. However, the epidemic curve was given. Therefore, the estimated reproduction number probably increased by extending the period of infectiousness from seven days to nine days.

Moreover, when using Poisson likelihood for evaluation, R_0_ alone was found to be significantly lower than in the base case. Although the difference was not large, it might engender some difference attributable to the selection of the evaluation function.

Among children and younger adults, the proportion of asymptomatic infected people might be larger than among elderly people, but that point remains uncertain. Lower susceptibility of children [3–5] might imply a higher proportion of asymptomatic cases in children. In this sense, the finding of 3/23 among elderly people might be the lower bound of the proportion for the general population. Overall, sensitivity analysis led to similar estimation with the base case. Even though significant difference was found through comparison with the base case only in R_0_, differences among them were not large, quantitatively speaking. Therefore, one might conclude that the obtained results were more or less robust.

The average incubation period applied in another study was 5.1 days [21], but it was 6.6 days for the present study. However, the latter number based on the epidemic curve considered in the present study, is certainly consistent with the data. It might reflect lower severity among patients in Japan than in other countries such as the US. The estimated value of the reproduction number probably would be smaller if we adopt a slightly shorter incubation period, which is a similar result to that found with sensitivity analysis for infectiousness before onset.

Many weeks passed since the end of the study period. Therefore, the reporting lag almost disappeared. If timely estimation, meaning estimation using data from a day prior, was necessary, then we used an adjusted delay for reported data [8, 22, 23].

We used the minimized sum of the absolute values of discrepancies for the bootstrapped epidemic curve and the fitted values, instead of the minimized sum of squared residuals such as maximum likelihood estimation based on a normal distribution. In general, minimization of the sum of the absolute values is more robust than minimization of the sum of squares because the absolute value is less sensitive than squared values to the effects of outliers [24–26]. The daily epidemic curve sometimes exhibits spikes according to the day of the week and for other reasons. Especially, few patients per day were reported during the early stage of the outbreak. Therefore, spikes might be quite large. These spikes are probably outliers. They might overly affect the estimator. For this reason, we prefer minimization of the sum of the absolute values to minimization of the sum of squared values when analyzing daily data in the earlier stage of the outbreak.

Moreover, we chose non-parametric approaches using actual data in preference to parametric approaches assuming a particular distribution. Such an assumption might affect results through miss-specification. Therefore, we prefer to use the actual distribution of incubation periods or epidemic curves with no unnecessary or restrictive assumption. Sensitivity analysis using minimum least squares of errors or using Poisson likelihood to maximize showed very similar results to those obtained with the base case. Although R_0_ and R_a_ were slightly higher, R_e_ were slightly lower. That result might indicate that some outlier in the pre-VECSC and post-VECSC period pulls the estimator in sensitivity analysis.

This study has some limitations. The first is that even though we evaluated VECSC, the respective effects of voluntary event cancellation and school closure cannot be discerned. To distinguish their respective effects, one would have to develop a model with several age classes. School closure mainly affects contact patterns among schoolchildren; voluntary event cancellation mainly affects patterns among adults. Therefore, studies of those respective age groups might elucidate the separate effects of these policies. That stands as a challenge for our future study.

The second point is underascertainment. Although the epidemic curves of COVID-19 in all countries are subject to underascertainment, it might be very difficult to evaluate the degree to which they are affected. It might bias the estimation result.

A third point is a lack of estimation of the outcomes such as dead or severe cases or necessary medical resources for the care of COVID-19 patients. We particularly examined how policies affect the reproduction number under countermeasures. Therefore, we ignored prediction of the entire course of the outbreak and its outcomes such as the number of deaths. Nevertheless, outcomes are expected to be a primary concern for modelling. Moreover, the collapse of medical services can be expected to engender worse outcomes even if the reproduction number remains unchanged. Prediction of the effects of severe policies including lockdowns is anticipated as a challenge to be addressed in our future research.

## Conclusion

Results have demonstrated that VECSC can reduce the infectiousness of COVID-19 considerably: approximately to one. However, the figure is probably greater than one. Outbreaks might continue for a long time. Therefore, lockdown policies are expected to be as effective as VECSC if they are executed carefully. After VECSC, the reproduction number escalated again beyond that before VECSC. Similar phenomena might be observed after a lockdown. It is our earnest hope that results of the present study can contribute to governmental policy-making related to lockdown measures or other countermeasures to combat the spread and destruction of COVID-19.

The present study was based on the authors’ opinions. Neither the results nor implications reflect any stance or policy of professionally affiliated bodies.

## Data Availability

Japan Ministry of Health, Labour and Welfare. Press Releases. (in Japanese) https://www.mhlw.go.jp/stf/houdou/houdou_list_202001.html.

## Acknowledgments

We acknowledge the great efforts of all staff at public health centers, medical institutions, and other facilities who are fighting the spread and destruction associated with COVID-19.

